# Temporal and Geographic Variation in Outcomes After Poor-Grade Aneurysmal Subarachnoid Hemorrhage: A Systematic Review and Meta-analysis

**DOI:** 10.64898/2026.06.29.26356892

**Authors:** Airton Leonardo De Olivera Manoel, Ali Msheik, Fernando G. Zampieri, Ruben Peralta, Ghaya Al Rumaihi, Hassan Al-Thani, Jose Ignacio Suarez

**Author notes:** Corresponding author: Airton Leonardo de Oliveira Manoel Trauma ICU, Trauma Surgery Section Hamad General Hospital, Hamad Medical Corporation, Doha, Qatar.

## Abstract

**Background:** Poor-grade aneurysmal subarachnoid hemorrhage (aSAH) remains associated with high mortality and severe disability, yet contemporary outcomes may differ substantially from historical estimates. We performed a systematic review and meta-analysis to evaluate long-term outcomes after poor-grade aSAH and assess temporal, geographic, and treatment-related factors associated with prognosis.

**Methods:** PubMed/MEDLINE, Embase, Cochrane Central, Scopus, and Google Scholar were searched from inception through March 2026. Studies enrolling consecutive adults with poor-grade aSAH (World Federation of Neurosurgical Societies grades IV–V, Hunt–Hess grades IV–V, or equivalent) reporting mortality and/or functional outcomes at ≥3 months were included. To minimize survivorship bias, studies excluding untreated patients or patients dying before aneurysm treatment were excluded. Random-effects meta-analyses of proportions were performed using generalized linear mixed models. Prespecified subgroup analyses and exploratory meta-regression analyses evaluated temporal, geographic, and treatment-related factors associated with outcomes.

**Results:** Forty-two studies including 7,726 patients from 16 countries across 4 continents were included. The pooled favorable functional outcome rate was 27.2% (95% CI, 23.9%–30.8%), whereas pooled overall mortality was 53.3% (95% CI, 49.0%–57.5%). Pre- and post-treatment mortality were 25.9% and 33.9%, respectively. Aneurysm treatment rate was 72.0% (95% CI, 65.6%–77.7%). Favorable outcomes improved over time from 13.5% (95% CI, 7.0%-24.3%) in the 1980s to 33.7% in the 1990s but plateaued thereafter. In exploratory meta-regression analyses, higher aneurysm treatment rates were independently associated with improved favorable functional outcome (0.134 log-odds increase per 10% increase in treatment rate; p = 0.01) and lower mortality (−0.224 log-odds per 10% increase in treatment rate; p < .001). Publication year was associated with lower mortality (p = 0.03) but not favorable outcome. Geographic region, country income group, and the proportion of grade V patients were not independently associated with outcomes.

**Conclusions:** Mortality after poor-grade aSAH remains high, but approximately one-third of patients achieved favorable outcome. Higher aneurysm treatment rates were independently associated with improved functional outcomes and lower mortality.

## Introduction

Aneurysmal subarachnoid hemorrhage (aSAH) remains a devastating neurological emergency associated with high mortality and long-term disability(1). Patients presenting with poor-grade aSAH, typically defined as World Federation of Neurosurgical Societies (WFNS) grades IV–V or Hunt–Hess (HH) grades IV–V, represent the most critically ill subgroup and historically have been considered to have extremely poor prognosis. Early studies from the pre-endovascular and neurocritical care eras reported extremely poor outcomes, with mortality rates approaching 70% among grade IV and nearly 100% in grade V patients(2). Importantly, these early treatment paradigms were largely developed for surgical risk assessment, and many patients with poor grade were managed conservatively. Consequently, these historical series likely contributed to therapeutic nihilism, frequent withholding of active treatment strategies, and premature withdrawal of life-sustaining therapies(3).

In population-based studies of aSAH, case-fatality has declined, although substantial regional variability exists(4). This reduction in mortality has coincided with the introduction of modern neurocritical care strategies, early aneurysm occlusion, endovascular therapy, improved delayed cerebral ischemia management, and the implementation of specialized high volume centers(5,6).

Although, the increasing recognition of the potential for meaningful neurological recovery in the poor-grade aSAH population has challenged historical assumptions of futility(7), the management of these patients has not advanced uniformly, and substantial hesitancy to offer aggressive treatment or to transfer to specialized neuroscience centers still persists(8). Nevertheless, reported outcomes remain highly heterogeneous across studies because of differences in patient selection, treatment aggressiveness, withdrawal of life-sustaining therapies practices, study epoch, and geographic variations. These factors introduce important confounding factors that may substantially influence observed outcomes and limit the interpretation of the existing literature(8,9).

More importantly, cohorts evaluating poor-grade aSAH outcomes have often been limited by the inclusion of highly selected patients, and the lack of representation of patients treated conservatively or patients who died before aneurysm treatment.

Therefore, we conducted a systematic review and meta-analysis to evaluate mortality and long-term functional outcomes after poor-grade aSAH, while specifically addressing survivorship bias through inclusion of studies reporting untreated patients and pre-treatment mortality. We additionally sought to evaluate temporal trends in outcomes across decades, explore geographic variations in outcomes, and assess potential study-level associations between outcomes of interest and publication year, proportion of grade V patients, aneurysm treatment rate, geographic region, and country income group. We hypothesized that functional outcome and mortality rates for aSAH patients have improved over time and across geographic regions.

## Methods

### Protocol and Reporting Standards

The research question and eligibility criteria were defined a priori. This systematic review and meta-analysis were conducted in accordance with the Preferred Reporting Items for Systematic Reviews and Meta-Analyses (PRISMA) guidelines and prospectively registered in the International Prospective Register of Systematic Reviews (PROSPERO; CRD420261358459).

### Eligibility criteria

Studies were eligible if they enrolled consecutive adult patients (≥18 years) with aneurysmal subarachnoid hemorrhage confirmed by computed tomography, CT angiography, digital subtraction angiography, or lumbar puncture and included patients classified as poor-grade aSAH, defined as World Federation of Neurosurgical Societies (WFNS) grades IV–V(10), Hunt–Hess grades IV–V(2), or equivalent severity scales. Eligible study designs included observational cohort studies and randomized controlled trials. Studies were required to report mortality and/or long-term functional outcomes assessed at ≥3 months using functional outcome scales such as the Glasgow Outcome Scale (GOS), GOS Extended (GOSE), modified Rankin Scale (mRS), or equivalent outcome scales, with extractable data specific to poor-grade patients. To minimize survivorship bias and ensure representation of the full disease spectrum, studies were included if they explicitly reported patients managed conservatively, patients who did not receive aneurysm treatment, and/or patients who died before aneurysm treatment. Studies including mixed-grade populations were eligible only when data for poor-grade patients could be clearly separated.

Studies were excluded if they lacked poor-grade–specific outcome data, excluded untreated patients or patients dying before aneurysm treatment without adequate reporting, reported only short-term outcomes (<3 months), or represented duplicate or overlapping cohorts. Reviews, editorials, conference abstracts, physiology-only studies, and feasibility studies without clinical outcome data were also excluded.

### Information Sources and Search Strategy

A comprehensive literature search was conducted in PubMed/MEDLINE, Embase, Cochrane Central Register of Controlled Trials (CENTRAL), Scopus, and Google Scholar from database inception through March 2026. Articles published in English were identified using combinations of the terms “aneurysmal subarachnoid hemorrhage,” “subarachnoid hemorrhage,” “poor-grade,” “high-grade,” “functional outcome,” “long-term functional outcome,” and “mortality.” The complete search strategy is available in the PROSPERO registration record (CRD420261358459). Additional studies were identified through forward citation searching, backward citation tracking of reference lists, and screening of the authors’ personal databases.

### Study Selection and Data Extraction

Two reviewers (A.L.O.M. and A.M.) independently screened titles and abstracts for eligibility. Full-text articles were subsequently reviewed when potentially eligible. Disagreements were resolved by consensus, with arbitration by a third reviewer (F.G.Z.) when necessary.

Study-level data were independently extracted by the two reviewers using a standardized data extraction form. Extracted variables included study characteristics (first author, publication year, country, study design, number of centers, and enrollment period), participant characteristics (sample size, grading system, proportion of grade IV and grade V patients), treatment characteristics (surgical clipping, endovascular coiling, conservative management, and reporting of pre-treatment deaths), and outcome variables (mortality, favorable and unfavorable functional outcome, outcome definition, follow-up duration, and neurological outcome scale used).

### Effect Measures

The primary denominator for all analyses was the total enrolled study population. Prespecified outcomes included favorable functional outcome, overall mortality, pre-treatment mortality, post-treatment mortality, aneurysm treatment rate, and treatment modality (surgical clipping, endovascular coiling, conservative management, or untreated status). Favorable functional outcome was defined according to each study’s reported criteria and most corresponded to GOS scores of 4–5 or mRS scores of 0–2 or 0–3. When multiple follow-up time points were reported, the longest available follow-up of at least 3 months was preferentially used for quantitative synthesis.

### Risk of Bias Assessment

Risk of bias was independently assessed by the two reviewers, with disagreements resolved by consensus and arbitration by a third reviewer when necessary. Observational studies were evaluated using the Newcastle–Ottawa Scale (NOS), and randomized controlled trials were assessed separately using the Cochrane Risk of Bias 2 (RoB 2) tool. Studies scoring ≥7 points on the NOS were classified as good quality, whereas studies scoring <7 points were classified as poor quality.

A formal assessment of certainty of evidence using the Grading of Recommendations Assessment, Development and Evaluation (GRADE) framework was not performed because this review primarily synthesized prognostic proportions derived from predominantly observational studies with substantial clinical and methodological heterogeneity.

### Statistical Analysis

Random-effects meta-analyses of proportions were performed for favorable functional outcome, overall mortality, pre-treatment mortality, post-treatment mortality among treated patients, and aneurysm treatment rate. Meta-analyses were fitted using generalized linear mixed models with logit-link transformation.

Crude event proportions, pooled proportions, 95% confidence intervals (CIs), 95% prediction intervals, between-study heterogeneity (I²), and between-study variance (τ²) were calculated. Values of I² of approximately 25%, 50%, and 75% were interpreted as representing low, moderate, and high heterogeneity, respectively.

Sensitivity analyses were performed using a two-step logit-transformed proportion approach with restricted maximum likelihood (REML) estimation and Hartung–Knapp adjustment. Funnel plots were generated to assess potential small-study effects and publication bias.

Exploratory subgroup analyses evaluated favorable functional outcome according to publication decade, grading system (WFNS versus Hunt–Hess), outcome definition, study design (prospective versus retrospective), center type (single-center versus multicenter), geographic region, and study quality. Geographic regions were categorized as Europe, North America, and Asia/Australia. Additional analyses evaluated outcomes according to World Bank fiscal year 2026 country income classification.

Exploratory random-effects meta-regression analyses were performed to examine associations between study-level characteristics and favorable functional outcome or overall mortality. Prespecified moderators included publication year, proportion of Grade V patients, aneurysm treatment rate, geographic region, and country income group. Because these analyses were conducted at the study level, results should be interpreted as ecological associations and should not be considered evidence of patient-level causal effects.

Treatment modality (surgical clipping versus endovascular coiling) and outcomes among conservatively managed patients were summarized descriptively because most studies did not report modality-specific outcome data suitable for quantitative synthesis.

All statistical analyses were performed using R (R Foundation for Statistical Computing, Vienna, Austria) with the metafor package.

### Reporting Bias Assessment

Potential reporting bias and small-study effects were explored visually using funnel plots for outcomes including at least 10 studies. Formal statistical testing for publication bias was interpreted cautiously because meta-analyses of single-arm proportions are particularly susceptible to substantial clinical and methodological heterogeneity. Efforts to minimize reporting bias included comprehensive database searches, forward and backward citation tracking, and screening of the authors’ personal databases.

## Results

### Study Selection and Characteristics

The study selection process is summarized in the PRISMA flow diagram (Figure 1). A total of 42 studies met the inclusion criteria and were included in the quantitative synthesis (Supplementary Table 1), comprising 41 observational cohorts studies [30 retrospective(7,11–39) and 11 prospective(40–50)] and 1 randomized controlled trial(51).

**Figure 01.**
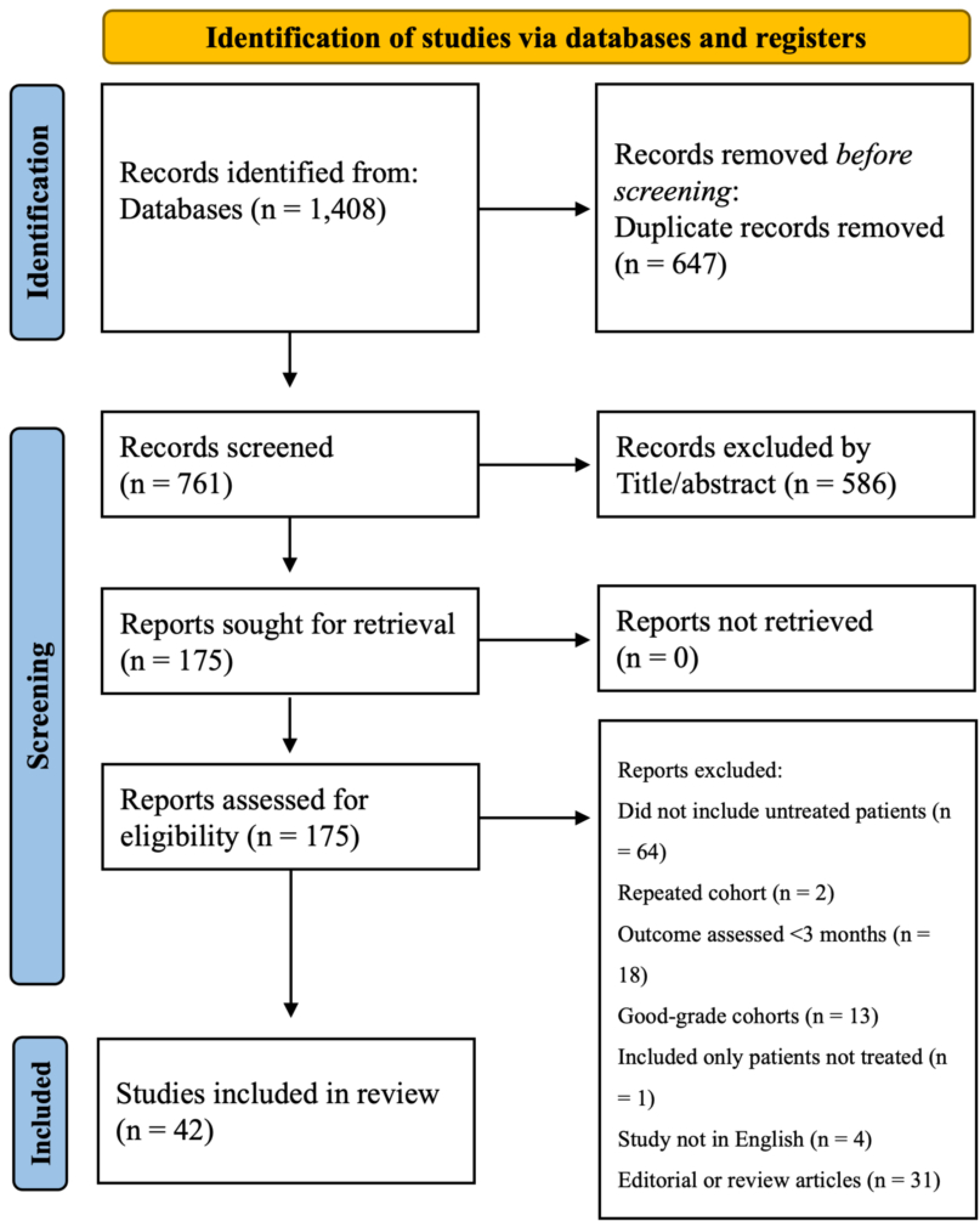
PRISMA Flow Diagram of Literature Search and Study Selection.

**Figure 02A.**
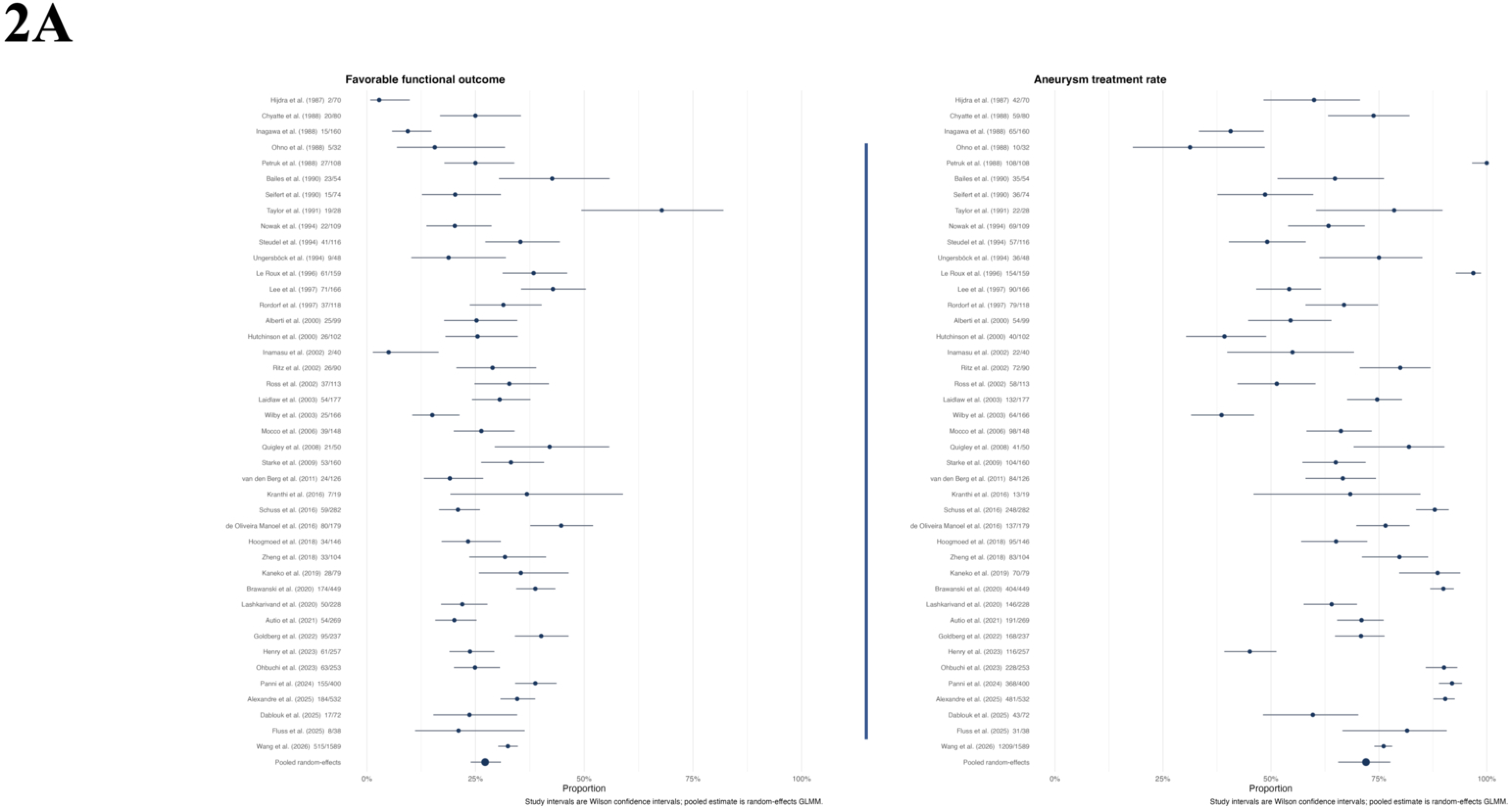
Forest plot of Favorable Outcome (left) and aneurysm treatment rate (right)

**Figure 02B.**
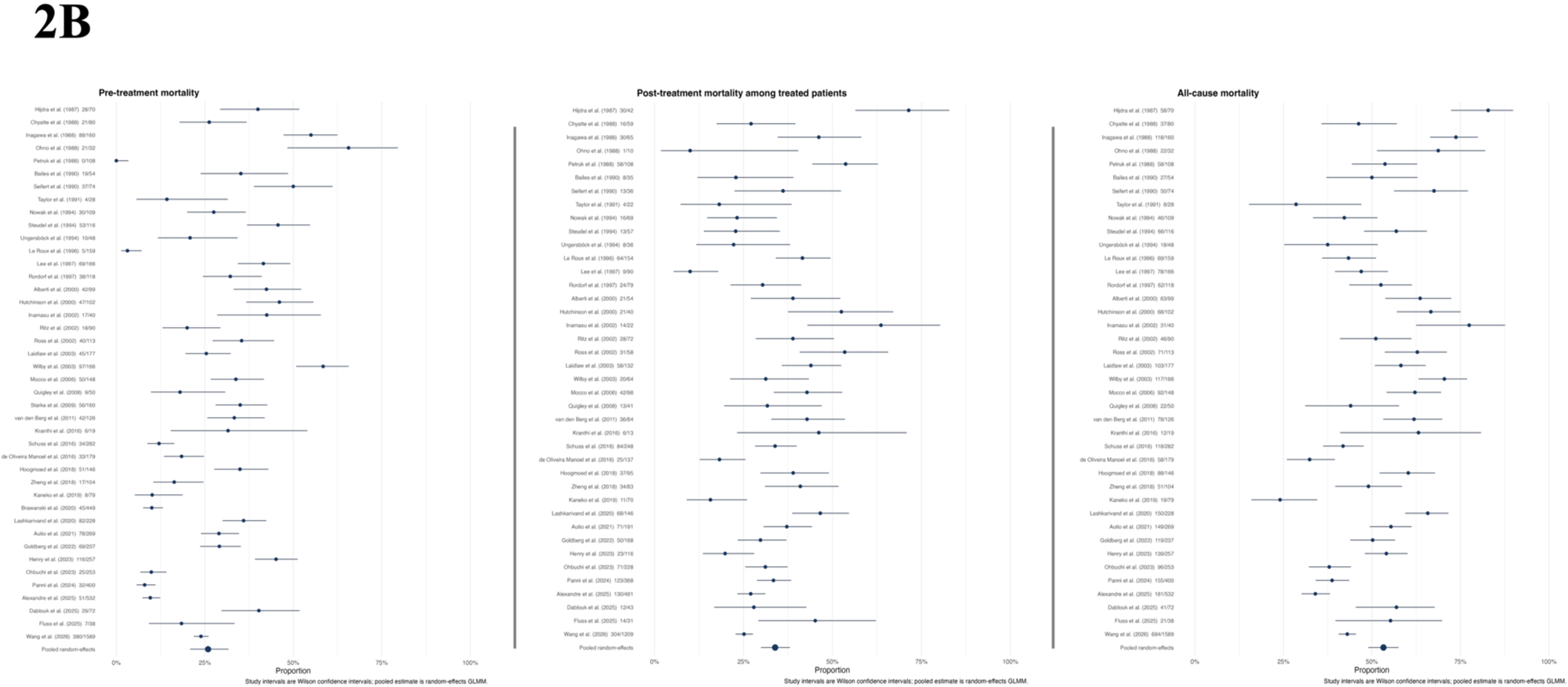
Forest plot of pre-treatment mortality (left), post-treatment mortality (center) and overall mortality (right)

The included studies were published between 1987 and 2026 and enrolled patients treated between 1977(41) and 2023(39), representing nearly five decades of clinical practice. Studies originated from 16 countries across four continents, including North America (United States, Canada), Europe (England, Scotland, Ireland, Germany, Switzerland, Italy, Netherlands, Norway, Finland), Asia (China, Japan, Korea, India), and Oceania (Australia). The largest contributions came from Germany (n=9), the United States (n=8), and Japan (n=5). One study was multinational, including centers from Germany and Switzerland.

Poor-grade aSAH was defined using the WFNS scale in 21 studies (50%) and the HH grading system in 19 studies (45.2%). Less frequently used grading systems included the Hunt and Kosnik classification and the Modified Botterell classification (1 study each, 2.4%).

Definitions of favorable functional outcome varied across studies and were most commonly based on GOS scores of 4–5 (20 studies), mRS scores of 0–2 (8 studies), or mRS scores of 0–3 (5 studies). A small number of studies used GOSE scores of 5–8, GOS score of 4, mRS scores of 0–1, or other author-defined criteria. Outcome assessment was most frequently performed at 3 months (14 studies) or 6 months (14 studies), followed by 12 months (8 studies). The longest reported follow-up durations were 26 months and 36 months.

### Risk of bias

Among the 41 observational studies, 16 (39.0%) were classified as good quality and 25 (61.0%) as poor quality according to the Newcastle–Ottawa Scale (Supplementary Figure 01). The single randomized controlled trial demonstrated some concerns for risk of bias according to the RoB 2 assessment, primarily related to deviations from intended interventions, missing outcome data, and selection of the reported results(51). Detailed risk-of-bias assessments are presented in Supplementary Tables 2A and 2B.

### Patient Characteristics

A total of 7,726 patients were included, of whom 3,232 (41.8%) were classified as grade IV and 3,879 (50.2%) as grade V. The remaining 615 patients (8.0%) lacked separate grade-specific reporting.

### Overall Outcomes

Table 1 and Figures 2A-B summarize the crude and pooled outcome estimates across the included studies. Favorable long-term functional outcome occurred in 2,316 of 7,726 patients (30.0%), corresponding to a pooled estimate of 27.2% (95% CI, 23.9%–30.8%). Overall mortality occurred in 3,489 patients (49.0%), with a pooled mortality estimate of 53.3% (95% CI, 49.0%–57.5%). Pre-treatment mortality accounted for 1,949 deaths (25.2%), whereas post-treatment mortality occurred in 1,641 of 5,154 patients (31.8%). The overall aneurysm treatment rate was 73.3%, corresponding to a pooled treatment estimate of 72.0% (95% CI, 65.6%–77.7%).

**Table 01.** Crude and Pooled Outcome Estimates Across Included Studies.

| **Outcome** | **Number of studies** | **Events** | **N** | **Crude** | **Pooled** | **95%CI** | **Prediction interval** | **I2(%)** | **tau^2** |
| --- | --- | --- | --- | --- | --- | --- | --- | --- | --- |
| **Favorable functional outcome** | 42 | 2316 | 7726 | 30.0% | 27.2% | 23.9% to 30.8% | 11.7% to 51.4% | 90.6 | 0.274 |
| **Overall mortality** | 40 | 3489 | 7117 | 49.0% | 53.3% | 49.0% to 57.5% | 29.2% to 76.0% | 91.3 | 0.263 |
| **Pre-treatment mortality** | 42 | 1949 | 7726 | 25.2% | 25.9% | 20.8% to 31.7% | 5.5% to 67.7% | 96.0 | 0.812 |
| **Post-treatment mortality** | 40 | 1641 | 5154 | 31.8% | 33.9% | 29.9% to 38.1% | 15.4% to 59.1% | 87.2 | 0.270 |
| **Aneurysm treatment rate** | 42 | 5662 | 7726 | 73.3% | 72.0% | 65.6% to 77.7% | 27.6% to 94.6% | 96.5 | 0.926 |

Considerable between-study heterogeneity was observed across all analyses, with I² values consistently exceeding 85%. Prediction intervals were wide for all major outcomes, indicating substantial variation in outcome estimates across different studies, treatment eras, and healthcare systems.

### Temporal Trends, Evolution of Treatment Strategies and Geographic Variation in Outcomes

Supplementary Figures 2–4 illustrate temporal trends in study inclusion, patient enrollment, and aneurysm treatment modalities across publication decades. Although the number of included studies remained relatively stable from the 1990s onward (Supplementary Figure 2), the number of included patients increased substantially over time, with more than half of all patients originating from studies published in the 2020s. Specifically, patient counts increased from 450 in the 1980s to 4,324 in the 2020s (Supplementary Figure 3).

Regarding aneurysm treatment modalities, earlier decades were dominated by surgical clipping, whereas endovascular coiling increased progressively beginning in the 2000s and became the predominant reported treatment modality in contemporary cohorts. By the 2020s, the number of reported coiling procedures exceeded clipping procedures, reflecting a major shift toward endovascular management in modern neurovascular practice (Supplementary Figure 4).

### Subgroup And Sensitivity Analyses

Subgroup analyses according to publication decade demonstrated substantial temporal variation in outcomes (Table 2A–C). Favorable functional outcome improved from 13.5% (95% CI 7.0–24.3%) in studies published during the 1980s to 33.7% (95% CI 25.7–42.8%) in the 1990s and remained relatively stable thereafter, ranging from 26.1% to 29.3% in subsequent decades (Figure 3). Conversely, pooled mortality decreased from 66.0% (95% CI 48.2–80.4%) in the 1980s to 48.3% (95% CI 42.2–54.6%) in the 1990s, increased transiently to 62.1% in the 2000s, and subsequently declined to 46.5% and 48.5% in studies published during the 2010s and 2020s, respectively. Over the same period, aneurysm treatment rates increased from 75.9% (95% CI 48.1–91.7%) in the 1980s to 78.6% (95% CI 68.9–86.1%) in the 2020s. Although treatment rates declined during the 1990s and 2000s, they subsequently increased, reaching their highest levels in the 2010s (77.8%) and 2020s (78.6%).

**Table 02.**
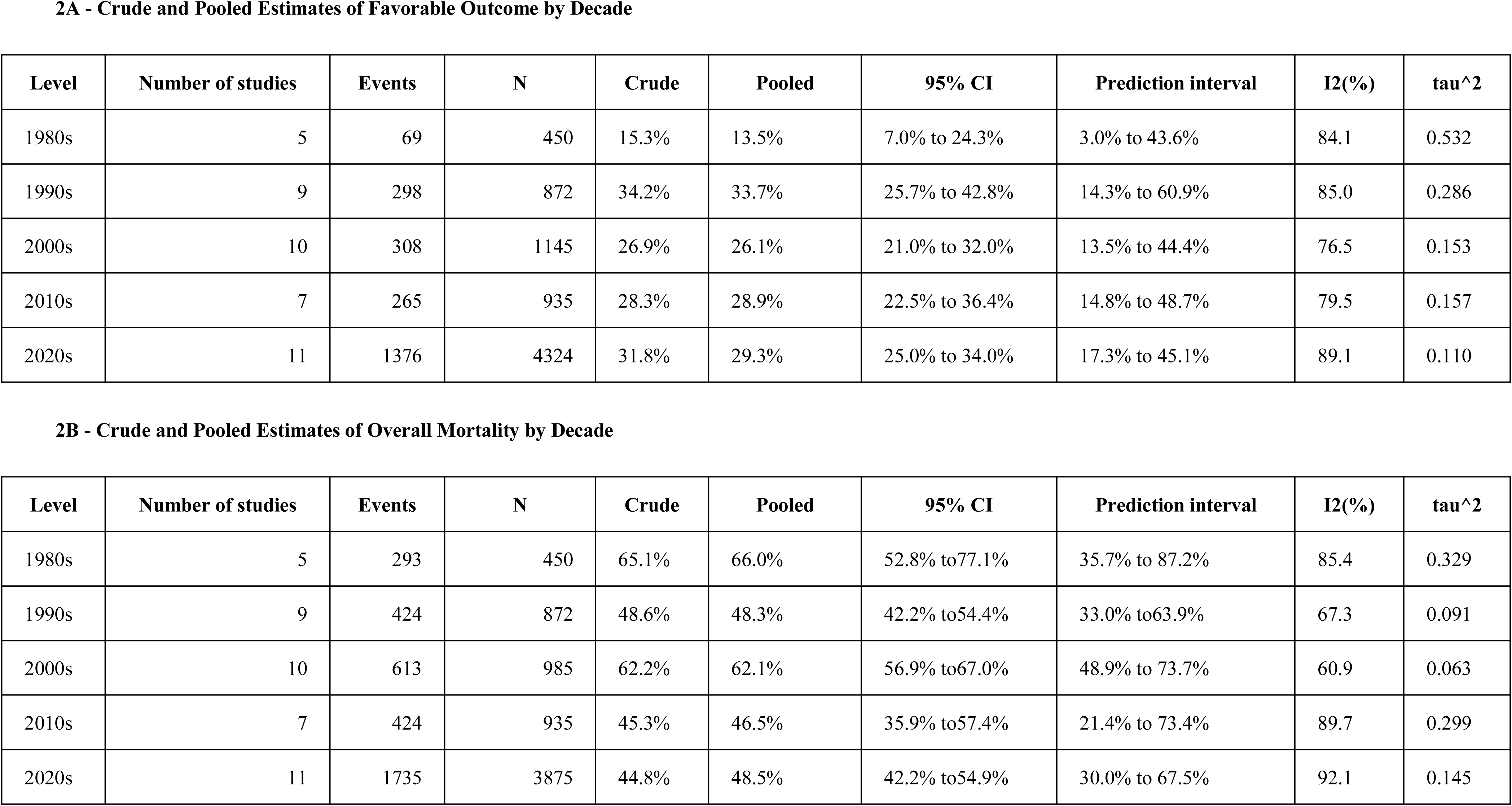
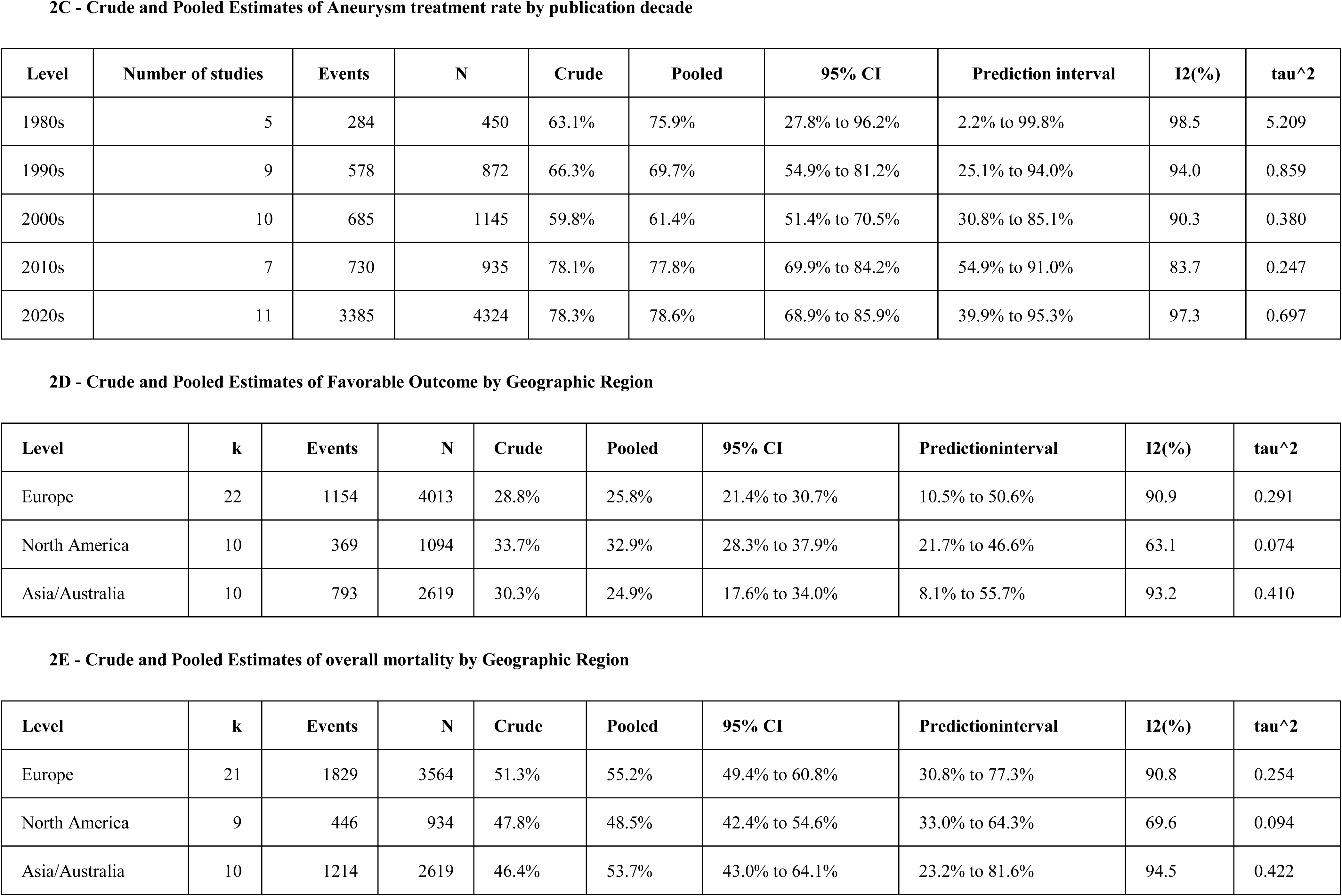
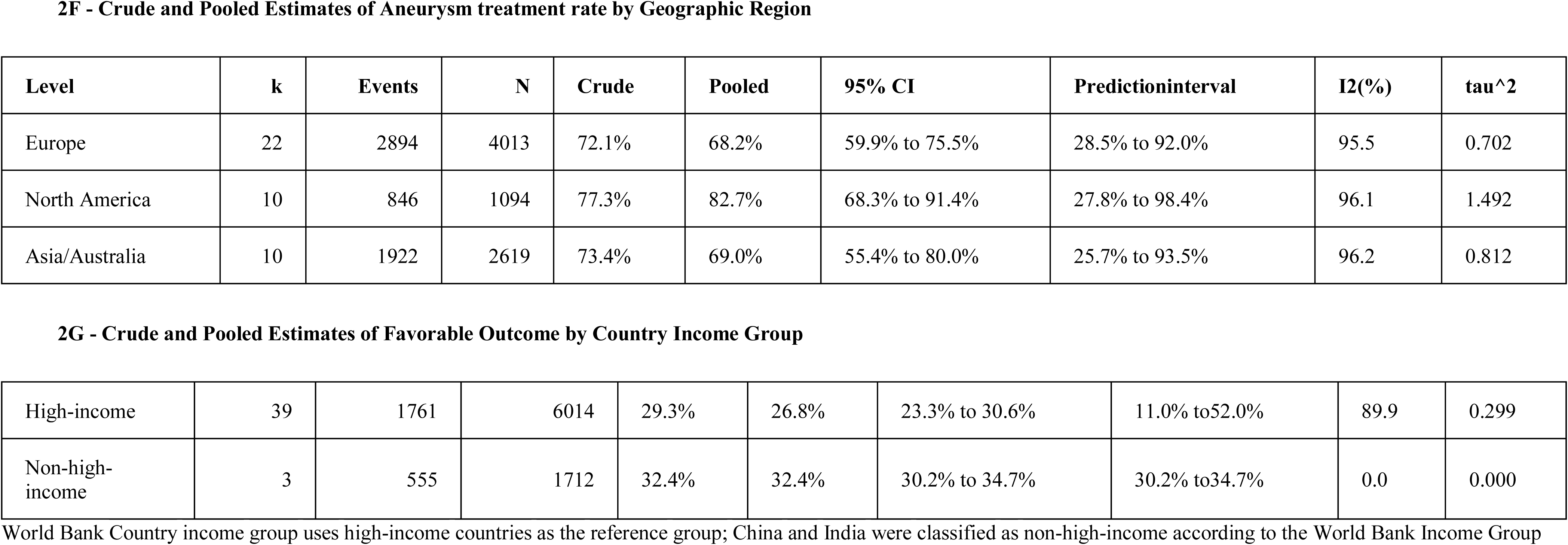
Subgroup and Sensitivity Analyses.

**Figures 3.**
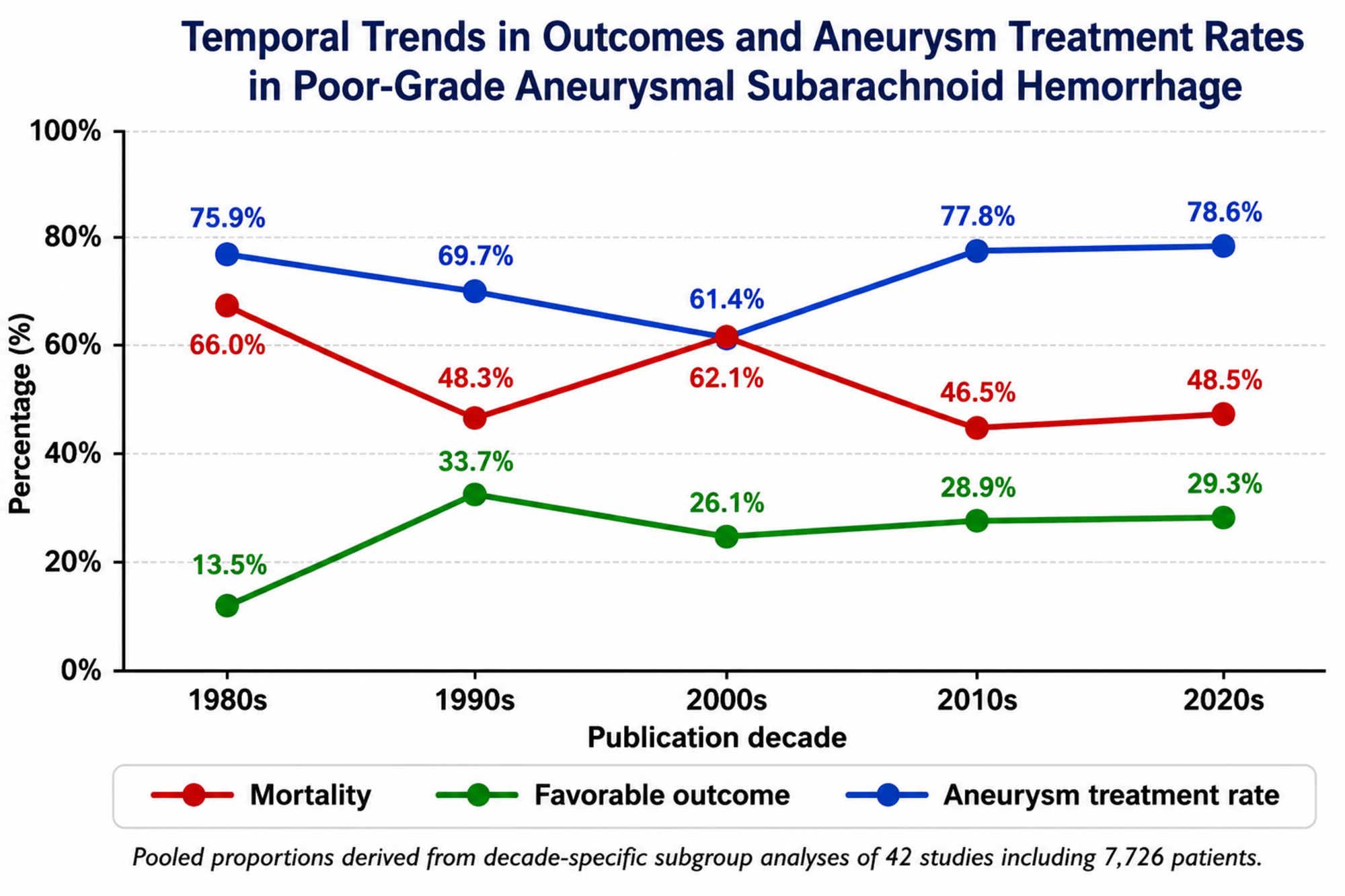
Temporal Trends in Outcomes and Aneurysm Treatment Rates in Poor-Grade Aneurysm Subarachnoid Hemorrhage.

Geographic subgroup analyses demonstrated modest regional variation in outcomes (Table 2D–F). North American studies reported the highest pooled favorable outcome rate (32.9%, 95% CI 28.3–37.9%) and the lowest pooled mortality rate (48.5%, 95% CI 42.4–54.6%), followed by European studies (favorable outcome 25.8%, [95% CI 21.4–30.7%]; mortality 49.4%, [95% CI 40.6–60.8%]) and studies from Asia/Australia (favorable outcome 24.9%, [95% CI 17.6–34.0%]; mortality 52.4%, [95% CI 43.5–61.1%]). Substantial heterogeneity persisted across regions, particularly among European and Asia/Australia studies (I² >90%), with wide prediction intervals indicating marked variability in outcomes across healthcare systems and treatment settings.

Subgroup analyses according to country income classification demonstrated broadly similar favorable outcome rates between high-income and non-high-income countries (Table 2G). Studies from high-income countries reported a pooled favorable outcome rate of 26.8% (95% CI 23.3–30.6%), whereas studies from non-high-income countries reported a pooled favorable outcome rate of 32.4% (95% CI 30.2–34.7%). However, this comparison should be interpreted cautiously because only three studies from two countries (China and India) were classified as non-high-income. Substantial heterogeneity was observed among studies from high-income countries (I² = 89.9%), whereas no statistical heterogeneity was detected in the limited non-high-income subgroup.

Additional subgroup and sensitivity analyses demonstrated generally consistent favorable functional outcome estimates across different grading systems, outcome definitions, study designs, center types, and study quality categories (Supplementary Tables 3–7).

### Exploratory Meta-regression Analysis

Table 03 and figure 04 summarizes the exploratory meta-regression analyses of study-level factors associated with favorable functional outcome and overall mortality.

**Table 03.** Exploratory Meta-regression Analyses of Study-Level Factors Associated with Favorable Functional Outcome and Overall Mortality.

| **Meta-regression Analyses of Factors Associated with Favorable Functional Outcome** |  |  |  |
| --- | --- | --- | --- |
| **Moderator** | **k** | **Estimate** | **p_value** |
| Publication year, per 10 years | 42 | 0.074 log-odds | 0.295 |
| Grade V proportion, per 10 percentage points | 40 | -0.030 log-odds | 0.500 |
| **Treatment rate, per 10 percentage points** | **42** | **0.134 log-odds** | **0.010** |
| Geographic region: North America vs Europe | 42 | 0.308 log-odds | 0.173 |
| Geographic region: Rest of world vs Europe | 42 | 0.005 log-odds | 0.984 |
| World Bank income group: Non-high-income vs High-income | 42 | 0.264 log-odds | 0.467 |
| **Meta-regression Analyses of Factors Associated with Overall Mortality** |  |  |  |
| **Publication year, per 10 years** | **40** | **-0.143 log-odds** | **0.032** |
| Grade V proportion, per 10 percentage points | 39 | 0.016 log-odds | 0.708 |
| **Treatment rate, per 10 percentage points** | **40** | **-0.224 log-odds** | **<0.001** |
| Geographic region: North America vs Europe | 40 | -0.259 log-odds | 0.263 |
| Geographic region: Rest of world vs Europe | 40 | -0.079 log-odds | 0.724 |

| Meta-regression Analyses of Factors Associated with Favorable Functional Outcome |  |  |  |
| --- | --- | --- | --- |
| Moderator | k | Estimate | p_value |
| World Bank income group: Non-high-income vs High-income | 40 | -0.147 log-odds | 0.679 |

**Figure 4.**
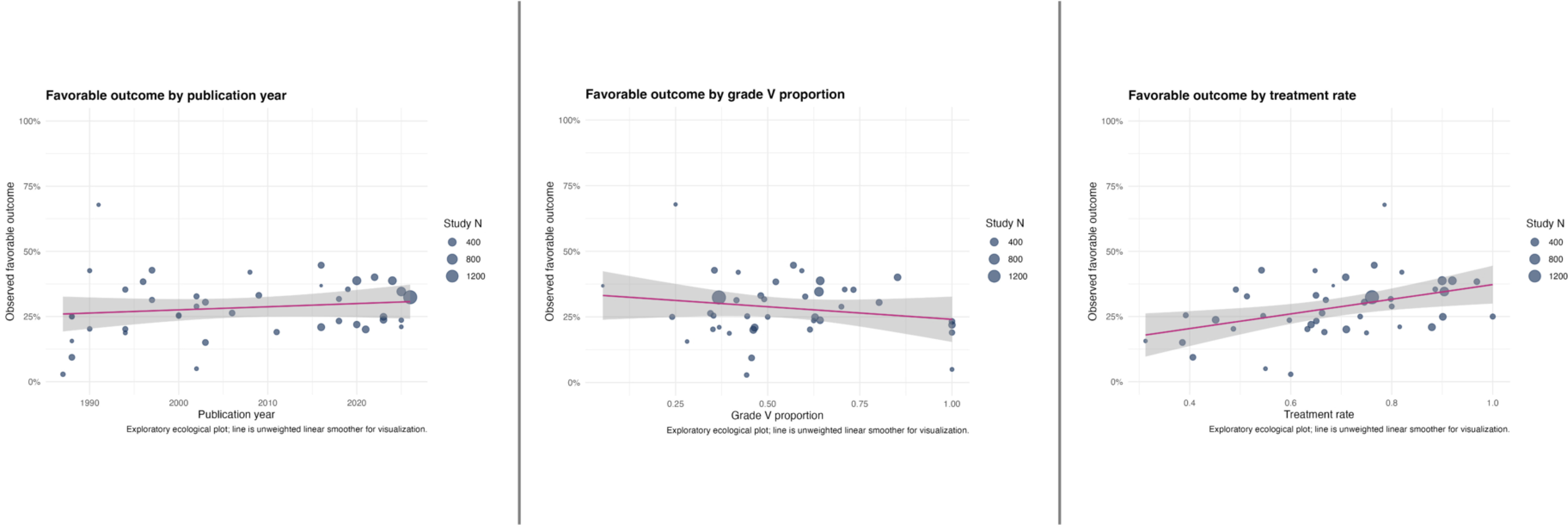
Exploratory Meta-regression Analysis.

Higher aneurysm treatment rates were significantly associated with improved favorable functional outcomes (estimate, 0.134 log-odds increase per 10% increase in treatment rate; *P* = 0.01). In contrast, publication year (estimate, 0.074 log-odds per decade; *P* = .30), proportion of grade V patients (estimate, −0.030 log-odds per 10% increase; *P* = .50), geographic region (North America vs Europe; estimate, 0.308 log-odds, p=0.173; and Asia/Australia vs Europe; estimate, 0.005 log-odds, p=0.984), and country income group (estimate, 0.264 log-odds, p=0.467) were not independently associated with favorable outcome.

Meta-regression analyses for overall mortality demonstrated that both publication year and aneurysm treatment rate were significantly associated with overall mortality. More recent studies reported lower mortality (estimate, −0.143 log-odds per decade; P=0.032), while higher aneurysm treatment rates were strongly associated with reduced mortality (estimate, −0.224 log-odds per 10% increase in treatment rate; P<0.001). In contrast, the proportion of grade V patients, geographic region, and country income group were not significantly associated with mortality.

### Assessment of Reporting Bias and Small-Study Effects

Visual inspection of funnel plots demonstrated mild-to-moderate asymmetry across several outcomes, suggesting the possibility of small-study effects, publication bias, selective reporting, and/or underlying clinical heterogeneity. Funnel plots for all-cause mortality (Supplementary Figure 5) and favorable functional outcome (Supplementary Figure 6) showed greater dispersion among smaller studies, with several small cohorts reporting more extreme outcome estimates.

Similar patterns were observed for pre-treatment mortality (Supplementary Figure 7), post-treatment mortality among treated patients (Supplementary Figure 8), and aneurysm treatment rates (Supplementary Figure 9).

Nevertheless, most studies were distributed within the expected pseudo–95% confidence limits, and the observed asymmetry should be interpreted cautiously given the substantial between-study heterogeneity and the predominance of observational studies.

Interpretation of funnel-plot asymmetry should be with caution because meta-analyses of single-arm proportions are particularly susceptible to distortion from substantial between-study heterogeneity, differences in treatment selection, variability in withdrawal-of-care practices, and evolving neurocritical care strategies across decades and geographic regions. Consequently, observed asymmetry may reflect true clinical and methodological heterogeneity rather than publication bias alone.

## Discussion

In this systematic review and meta-analysis of 42 studies including 7,726 unselected patients with poor-grade aSAH from 16 countries across four continents, overall mortality was high, exceeding 50%. Nevertheless, approximately one-third of patients achieved favorable long-term functional outcomes. Outcomes improved substantially after the 1990s and subsequently remained relatively stable despite continuing advances in neurovascular and neurocritical care. Among the study-level factors evaluated, higher aneurysm treatment rates were the only variable independently associated with both improved favorable outcomes and lower mortality. Collectively, these findings suggest that aggressive aneurysm treatment may be an important determinant of prognosis in poor-grade aSAH and challenge the traditional perception that severe neurological presentation invariably predicts futility(5,20).

Through our search strategy, we identified numerous studies reporting outcomes in poor-grade aSAH; however, many excluded important patient subgroups, particularly conservatively managed patients and those who died before aneurysm treatment. Such exclusions may substantially overestimate favorable outcomes because only patients surviving long enough to receive intervention are included in the analysis. Conversely, studies reporting only short-term outcomes may underestimate eventual neurological recovery. To minimize these limitations, our study preferentially included unselected poor-grade cohorts whenever possible, incorporating conservatively managed patients and pre-treatment mortality to better represent the full spectrum of disease severity encountered in clinical practice.

### Temporal Evolution of Outcomes and Treatment Strategies

Over the past five decades, outcomes after poor-grade aSAH have evolved(7). Earlier studies commonly used admission neurological grade to determine eligibility for aneurysm treatment, resulting in exclusion of many severely affected patients from neurosurgical intervention, which resulted in extremely poor outcomes and reinforced a perception of therapeutic futility in this population(2). Later studies treating patient more aggressively suggested that conservative management of poor-grade patients could increase the risk of rebleeding(28), and also lead to withdrawal of life-sustaining therapy in approximately one-third of patients who might otherwise achieve meaningful functional recovery if treated aggressively(20).

Our findings demonstrate substantial temporal evolution in both management strategies and outcomes. Earlier studies from the 1980s predominantly reflected highly selective surgical cohorts managed during the pre-endovascular era and reported very low rates of favorable recovery(11–13,41,51). In contrast, contemporary studies included substantially larger patient populations, broader use of endovascular therapy, and higher aneurysm treatment rates, coinciding with improved functional outcomes over time(7,50). Favorable functional outcome increased from 13.5% in the 1980s to approximately 30% thereafter, while mortality declined substantially compared with the earliest cohorts.

Several factors may have contributed to these improvements, including advances in diagnostic imaging(52), the introduction of nimodipine(51), developments in microsurgical and endovascular techniques, improved management of delayed cerebral ischemia, enhanced intracranial pressure monitoring and treatment(5), specialized neurovascular intensive care units(53), and regionalization of care to high-volume centers(54). However, despite these advances, favorable outcomes appear to have plateaued since the 1990s. Interestingly, publication year itself was not independently associated with favorable outcome in meta-regression analyses, whereas aneurysm treatment rate remained significantly associated with both favorable outcome and mortality. These findings suggest that increasing treatment intensity may explain a substantial proportion of the temporal improvement observed across studies.

### Exploratory Ecological Associations

Exploratory meta-regression analyses demonstrated substantial variability in outcomes across studies. Studies with higher aneurysm treatment rates consistently reported both higher favorable outcome rates and lower mortality. In contrast, studies including greater proportions of grade V patients tended to report somewhat worse outcomes(28), although this relationship was weak and statistically nonsignificant.

These findings suggest that differences in treatment aggressiveness pursuing aneurysm occlusion may contribute more to outcome variability than differences in baseline neurological grade alone. However, these analyses should be interpreted cautiously because they are based on study-level data and therefore represent ecological associations rather than patient-level causal effects. Although the observed associations are biologically plausible and clinically consistent, they cannot establish that increasing treatment rates directly improves outcomes in individual patients.

### Conservative Management and Outcome of Untreated Patients

Only a limited number of studies in our meta-analysis explicitly reported outcomes among conservatively managed or untreated patients(25,27,46,50). Across these studies, prognosis was consistently extremely poor. Wang et al.(50) included 380 patients receiving conservative treatment, representing 23.9% of their corhot, of whom only 9.7% were alive at 3 years. Conservative management was independently associated with mortality in their multivariable analyses. Similarly, Ritz et al.(25), Mocco et al.(46), and Starke et al(27). reported mortality rates approaching 100% among untreated patients.

More recently, Yang et al.(55) evaluated the natural history of untreated ruptured aneurysms using data from the Chinese Multicenter Cerebral Aneurysm Database. Among 941 untreated patients, 58.6% died within the first month after symptom onset and 68.1% died within 2 years. Poor Hunt–Hess grade, loss of consciousness at presentation, and aneurysm size ≥5 mm were independently associated with increased mortality.

Importantly, inconsistent reporting and frequent exclusion of untreated patients represent major methodological limitations within the poor-grade aSAH literature. Many studies excluded patients who died before aneurysm occlusion or were never considered candidates for aggressive intervention, thereby introducing substantial survivorship and selection bias. Consequently, studies restricted to treated patients may substantially overestimate favorable outcomes within the overall poor-grade population. By incorporating conservatively managed patients and pre-treatment mortality whenever possible, our meta-analysis provides a more representative estimate of prognosis across the entire spectrum of poor-grade aSAH.

### Grade-Specific Outcomes

Only three studies provided outcome data stratified according to grade IV versus grade V presentation(27,29,46). Among these studies, outcomes were consistently better among grade IV patients. Mocco et al. reported favorable outcomes in 62.2% of grade IV patients compared with 20.8% of grade V patients(46). Similarly, Starke et al.(27) reported favorable outcomes in 53.0% versus 11.7%, respectively, while Schuss et al.(29) reported favorable outcomes in 33.1% of WFNS grade IV patients compared with 9.3% of grade V patients.

Despite these differences, our meta-regression analyses demonstrated that the proportion of grade V patients was not independently associated with favorable outcome or mortality. Although this finding should not be interpreted as evidence that neurological grade lacks prognostic significance, it suggests that differences in admission grade alone do not explain the substantial heterogeneity observed across studies. Other factors such as treatment intensity, patient selection, timing of intervention, and healthcare system characteristics may play equally important roles in determining outcome.

### Treatment Modality and Outcome

We were unable to perform a formal meta-analysis comparing clipping and coiling because most studies did not provide modality-specific outcomes. Nevertheless, the limited available data provide several important observations.

Van den Berg et al.(28) reported higher favorable outcome rates among surgically treated patients than among patients undergoing endovascular coiling, whereas Mocco et al.(46) found no significant differences in survival or 12-month functional outcomes between modalities. In contrast, Wang et al., which contributed the largest cohort to the present review, reported lower mortality and superior overall efficacy among patients treated with endovascular techniques compared with surgical clipping(50).

Treatment patterns evolved substantially over time. Earlier decades were dominated by surgical clipping, whereas endovascular coiling increased progressively beginning in the 2000s and became the predominant reported treatment modality in contemporary studies. By the 2020s, coiling procedures exceeded clipping procedures, reflecting the profound shift toward endovascular management observed throughout modern neurovascular practice. However, the current literature remains insufficient to determine whether one modality confers superior outcomes specifically among poor-grade patients, highlighting an important area for future research.

### Regional Variability in Outcomes

Our subgroup analyses demonstrated modest regional variation in outcomes. North American studies reported the highest pooled favorable outcome rates and lowest mortality, whereas European and Asia/Australia studies reported somewhat less favorable results. However, substantial heterogeneity was observed within all geographic regions, indicating that regional differences likely reflect variations in patient selection, treatment practices, healthcare organization, and study methodology rather than geographic location itself.

These findings are broadly consistent with previous epidemiological studies demonstrating substantial worldwide variability in aSAH outcomes(1,4). Differences in access to specialized neurovascular centers, timing of aneurysm treatment, neurocritical care resources, referral pathways, and implementation of aggressive management strategies may partially explain these variations. Importantly, geographic region was not independently associated with favorable outcome or mortality in meta-regression analyses, suggesting that organizational and treatment-related factors may be more important determinants of outcome than geography alone.

Similarly, analyses according to country income classification demonstrated no significant association between income group and outcome. However, only three studies originated from non-high-income countries, limiting interpretation.

### Future Perspectives and Research Implications

Despite substantial advances in neurovascular and neurocritical care, important gaps remain in the management and prognostication of poor-grade aSAH. The current literature consists predominantly of retrospective observational studies with considerable heterogeneity in patient selection, treatment thresholds, outcome definitions, and follow-up duration. Future research should prioritize large prospective multicenter registries and collaborative international datasets using standardized data collection methodologies and harmonized outcome definitions.

One of the major limitations identified in this meta-analysis was the inconsistent inclusion and reporting of untreated patients, pre-treatment mortality, and withdrawal-of-care decisions. Future studies should systematically report the entire clinical trajectory of poor-grade patients, including reasons for treatment limitation and withdrawal of life-sustaining therapies. Such reporting is essential to minimize survivorship bias and address the recognized risk of self-fulfilling prophecy in poor-grade aSAH research.

The implementation of standardized Common Data Elements (CDEs) for aSAH may substantially improve future research quality and comparability across studies(56). Standardized use of these elements would facilitate individual patient-data meta-analyses, external validation of prognostic models, and more robust multicenter collaborative investigations.

Another major limitation of the current literature is the reliance on dichotomized functional outcome measures such as the modified Rankin Scale and Glasgow Outcome Scale. Increasing evidence demonstrates that many survivors experience substantial cognitive, emotional, behavioral, and quality-of-life impairments despite apparently favorable functional recovery(57). Future studies should therefore incorporate neuropsychological assessments, return-to-work measures, emotional outcomes, caregiver burden, and health-related quality-of-life metrics.

Additional research is also needed to better characterize the heterogeneity within poor-grade aSAH itself. Patients classified as grade IV and grade V likely represent biologically distinct populations with markedly different recovery potential. Future studies incorporating advanced imaging biomarkers, physiological monitoring, multimodal neuromonitoring, and longitudinal neurological trajectories may improve individualized prognostication and patient selection for aggressive intervention.

Finally, although increasing aneurysm treatment rates were associated with improved outcomes in our analyses, the optimal management strategy remains incompletely defined. High-quality comparative effectiveness studies evaluating timing of aneurysm occlusion, surgical versus endovascular approaches, decompressive craniectomy, cerebrospinal fluid diversion, delayed cerebral ischemia management, and advanced neurocritical care interventions remain urgently needed.

### Limitations

Several limitations should be considered when interpreting our findings. First, the evidence base consisted predominantly of observational studies, most of which were retrospective and therefore susceptible to selection bias, residual confounding, and incomplete outcome reporting. Second, substantial between-study heterogeneity was observed across nearly all analyses, likely reflecting differences in patient selection, treatment strategies, outcome definitions, follow-up duration, and healthcare systems. Third, favorable functional outcome was variably defined across studies, most commonly using GOS 4–5, mRS 0–2, or mRS 0–3, which may have influenced pooled estimates. Fourth, reporting of treatment limitation decisions and withdrawal of life-sustaining therapies was inconsistent. Fifth, only a minority of studies reported grade-specific or treatment modality–specific outcomes, precluding formal quantitative synthesis of these clinically important questions. Finally, the meta-regression analyses were exploratory and based on study-level rather than individual patient-level data; therefore, the observed associations should be interpreted as ecological and hypothesis-generating rather than causal.

## Conclusion

In this systematic review and meta-analysis of poor-grade aSAH, mortality exceeded 50%, yet approximately one-third of patients achieved favorable long-term functional outcomes. Outcomes improved substantially after the 1990s and have remained relatively stable in contemporary cohorts. Higher aneurysm treatment rates were the only study-level factor independently associated with both improved functional outcomes and lower mortality, supporting the potential value of aggressive aneurysm management in appropriately selected patients. These findings challenge the traditional perception that poor-grade presentation uniformly predicts futility and suggest that meaningful recovery remains achievable in a substantial proportion of patients. Future prospective multicenter studies using standardized outcome measures and comprehensive long-term follow-up are needed to refine prognostication and optimize treatment strategies for this high-risk population.

## Data Availability

Extracted study-level data, analytic code, and supplementary materials are available from the corresponding author upon reasonable request.

## Declarations

## Author Contributions

A.L.O.M. conceived, designed the study, and drafted the manuscript. A.L.O.M. and A.M. performed study screening, data extraction, and risk-of-bias assessment. F.G.Z. adjudicated disagreements during study selection and data extraction. F.G.Z. performed the statistical analyses. A.M., F.G.Z., R.P., G.A.R., H.A.-T., and J.I.S. contributed to data interpretation and critically revised the manuscript for important intellectual content. All authors reviewed and approved the final manuscript and agree to be accountable for all aspects of the work.

## SUPPLEMENTARY ONLINE MATERIAL

**Supplementary Table 1 - Characteristics of Included Studies and Patient Populations**

**Supplementary Table 2A. Risk of Bias Assessment of Included Cohort Studies Using the Newcastle–Ottawa Scale**

**Supplementary Table 2B. Risk of Bias Assessment of Included Randomized Clinical Trial Using the RoB 2 Tool**

## ADDITIONAL SUBGROUP AND SENSITIVITY ANALYSES

**Supplementary Table 3 - Favorable outcome by grading system**

**Supplementary Table 4 - Favorable outcome by outcome definition**

**Supplementary Table 5 - Favorable outcome by study design timing**

**Supplementary Table 6 - Favorable outcome by center type**

**Supplementary Table 7 - Favorable outcome by quality group**

## FUNNEL PLOTS

**Supplementary Figure 01 – Risk of Bias and Quality Assessment Summary**

**Supplementary Figure 02 – Included Studies by Publication Decade**

**Supplementary Figure 03 - Included Patients by Publication Decade**

**Supplementary Figure 04 – Reported Aneurysm Treatment Modality by Decade**

**Supplemental Figure 05 - Funnel Plot: All-Cause of Mortality**

**Supplemental Figure 06 - Funnel Plot: Favorable Outcome**

**Supplemental Figure 07 - Funnel Plot: Pre-treatment Mortality**

**Supplemental Figure 08 - Funnel Plot: Post-treatment Mortality**

**Supplemental Figure 09 - Funnel Plot: Treatment Rate**

## References

1. Feigin VL, Abate MD, Abate YH, Abd ElHafeez S, Abd-Allah F, Abdelalim A, et al. Global, regional, and national burden of stroke and its risk factors, 1990–2021: a systematic analysis for the Global Burden of Disease Study 2021. Lancet Neurol. 2024 Oct;23(10):973–1003. doi:10.1016/S1474-4422(24)00369-7

2. Hunt WE, Hess RM. Surgical Risk as Related to Time of Intervention in the Repair of Intracranial Aneurysms. J Neurosurg. 1968 Jan;28(1):14–20. doi:10.3171/jns.1968.28.1.0014

3. Hoogmoed J, De Oliveira Manoel AL, Coert BA, Marotta TR, Macdonald RL, Vandertop WP, et al. Why Do Patients with Poor-Grade Subarachnoid Hemorrhage Die? World Neurosurg. 2019 Nov;131:e508–13. doi:10.1016/j.wneu.2019.07.221

4. Nieuwkamp DJ, Setz LE, Algra A, Linn FH, De Rooij NK, Rinkel GJ. Changes in case fatality of aneurysmal subarachnoid haemorrhage over time, according to age, sex, and region: a meta-analysis. Lancet Neurol. 2009 Jul;8(7):635–42. doi:10.1016/S1474-4422(09)70126-7

5. De Oliveira Manoel AL, Goffi A, Marotta TR, Schweizer TA, Abrahamson S, Macdonald RL. The critical care management of poor-grade subarachnoid haemorrhage. Crit Care. 2016 Dec;20(1):21. doi:10.1186/s13054-016-1193-9

6. McNeill L, English SW, Borg N, Matta BF, Menon DK. Effects of Institutional Caseload of Subarachnoid Hemorrhage on Mortality: A Secondary Analysis of Administrative Data. Stroke. 2013 Mar;44(3):647–52. doi:10.1161/STROKEAHA.112.681254

7. De Oliveira Manoel AL, Mansur A, Silva GS, Germans MR, Jaja BNR, Kouzmina E, et al. Functional Outcome After Poor-Grade Subarachnoid Hemorrhage: A Single-Center Study and Systematic Literature Review. Neurocrit Care. 2016 Dec;25(3):338–50. doi:10.1007/s12028-016-0305-3

8. Hoogmoed J, Van Den Berg R, Coert BA, Rinkel GJE, Vandertop WP, Verbaan D. A strategy to expeditious invasive treatment improves clinical outcome in comatose patients with aneurysmal subarachnoid haemorrhage. Eur J Neurol. 2017 Jan;24(1):82–9. doi:10.1111/ene.13134

9. Al-Tamimi YZ. Management of poor-grade subarachnoid haemorrhage: a self-fulfilling prophecy of good outcome? Eur J Neurol. 2017 Jan;24(1):3–4. doi:10.1111/ene.13192

10. Teasdale GM, Drake CG, Hunt W, Kassell N, Sano K, Pertuiset B, et al. A universal subarachnoid hemorrhage scale: report of a committee of the World Federation of Neurosurgical Societies. J Neurol Neurosurg Psychiatry. 1988 Nov;51(11):1457–1457. doi:10.1136/jnnp.51.11.1457 PubMed PMID: 3236024; PubMed Central PMCID: PMC1032822.

11. Ohno K, Suzuki R, Masaoka H, Monma S, Matsushima Y, Inaba Y. A review of 102 consecutive patients with intracranial aneurysms in a community hospital in Japan. Acta Neurochir (Wien). 1988 Mar;94(1–2):23–7. doi:10.1007/BF01406610

12. Chyatte D, Fode NC, Sundt TM. Early versus late intracranial aneurysm surgery in subarachnoid hemorrhage. J Neurosurg. 1988 Sep;69(3):326–31. doi:10.3171/jns.1988.69.3.0326

13. Inagawa T, Takahashi M, Aoki H, Ishikawa S, Yoshimoto H. Aneurysmal subarachnoid hemorrhage in Izumo City and Shimane Prefecture of Japan. Outcome. Stroke. 1988 Feb;19(2):176–80. doi:10.1161/01.str.19.2.176

14. Bailes JE, Spetzler RF, Hadley MN, Baldwin HZ. Management morbidity and mortality of poor-grade aneurysm patients. J Neurosurg. 1990 Apr;72(4):559–66. doi:10.3171/jns.1990.72.4.0559

15. Seifert V, Trost HA, Stolke D. Management morbidity and mortality in grade IV and V patients with aneurysmal subarachnoid haemorrhage. Acta Neurochir (Wien). 1990;103(1–2):5–10. PubMed PMID: 2360467.

16. Taylor B, Harries P, Bullock R. Factors affecting outcome after surgery for intracranial aneurysm in Glasgow. Br J Neurosurg. 1991;5(6):591–600. PubMed PMID: 1772605.

17. Nowak G, Schwachenwald R, Arnold H. Early management in poor grade aneurysm patients. Acta Neurochir (Wien). 1994 Jan;126(1):33–7. doi:10.1007/bf01476491 PubMed PMID: 8154319.

18. Ungersbdck K, Bdcher-Schwarz H, Ulrich P, Wild A, Perneczky A. Aneurysm surgery of patients in poor grade condition. Indications and experience. Neurol Res. 1994;16(1):31–4. doi:10.1080/01616412.1994.11740188 PubMed PMID: 7913527.

19. Steudel WI, Reif J, Voges M. Modulated surgery in the management of ruptured intracranial aneurysm in poor grade patients. Neurol Res. 1994 Feb;16(1):49–53. doi:10.1080/01616412.1994.11740192

20. Le Roux PD, Elliott JP, Newell DW, Grady MS, Winn HR. Predicting outcome in poor-grade patients with subarachnoid hemorrhage: a retrospective review of 159 aggressively managed cases. J Neurosurg. 1996 Jul;85(1):39–49. doi:10.3171/jns.1996.85.1.0039

21. Lee KC, Huh SK, Park HS, Shin YS, Lee KS. Management of Poor-grade Patients with Ruptured Intracranial Aneurysm. Keio J Med. 1997;46(2):69–73. doi:10.2302/kjm.46.69

22. Rordorf G, Ogilvy CS, Gress DR, Crowell RM, Choi IS. Patients in poor neurological condition after subarachnoid hemorrhage: Early management and long-term outcome. Acta Neurochir (Wien). 1997 Dec;139(12):1143–51. doi:10.1007/BF01410974

23. Alberti O, Becker R, Benes L, Wallenfang T, Bertalanffy H. Initial hyperglycemia as an indicator of severity of the ictus in poor-grade patients with spontaneous subarachnoid hemorrhage. Clin Neurol Neurosurg. 2000 Jun;102(2):78–83. doi:10.1016/S0303-8467(00)00067-6

24. Inamasu J, Saito R, Mayanagi K. Endovascular Treatment for Poorest-grade Subarachnoid Hemorrhage in the Acute Stage: Has the Outcome Been Improved? Vol. 50. 2002;50(6).

25. Ritz R, Schwerdtfeger K, Strowitzki M, Donauer E, Koenig J, Steudel WI. Prognostic value of SSEP in early aneurysm surgery after SAH in poor-grade patients. Neurol Res. 2002 Dec;24(8):756–64. doi:10.1179/016164102101200852 PubMed PMID: 12500697.

26. Quigley MR, Salary M. Defining survivorship after high-grade aneurysmal subarachnoid hemorrhage. Surg Neurol. 2008 Mar;69(3):261–5. doi:10.1016/j.surneu.2007.02.013

27. Starke RM, Komotar RJ, Kim GH, Kellner CP, Otten ML, Hahn DK, et al. Evaluation of a revised Glasgow Coma Score scale in predicting long-term outcome of poor grade aneurysmal subarachnoid hemorrhage patients. J Clin Neurosci. 2009 Jul;16(7):894–9. doi:10.1016/j.jocn.2008.10.010 PubMed PMID: 19375327.

28. Van Den Berg R, Foumani M, Schröder RD, Peerdeman SM, Horn J, Bipat S, et al. Predictors of outcome in World Federation of Neurologic Surgeons grade V aneurysmal subarachnoid hemorrhage patients*: Crit Care Med. 2011 Dec;39(12):2722–7. doi:10.1097/CCM.0b013e3182282a70

29. Schuss P, Hadjiathanasiou A, Borger V, Wispel C, Vatter H, Güresir E. Poor-Grade Aneurysmal Subarachnoid Hemorrhage: Factors Influencing Functional Outcome—A Single-Center Series. World Neurosurg. 2016 Jan;85:125–9. doi:10.1016/j.wneu.2015.08.046

30. Hoogmoed J, Coert BA, Van Den Berg R, Roos YBWEM, Horn J, Vandertop WP, et al. Early Treatment Decisions in Poor-Grade Patients with Subarachnoid Hemorrhage. World Neurosurg. 2018 Nov;119:e568–73. doi:10.1016/j.wneu.2018.07.212

31. Kaneko J, Tagami T, Unemoto K, Tanaka C, Kuwamoto K, Sato S, et al. Functional Outcome Following Ultra-Early Treatment for Ruptured Aneurysms in Patients with Poor-Grade Subarachnoid Hemorrhage. J Nippon Med Sch. 2019;86(2):81–90. doi:10.1272/jnms.jnms.2019_86-203 PubMed PMID: 31130569.

32. Lashkarivand A, Sorteberg W, Rosseland LA, Sorteberg A. Survival and outcome in patients with aneurysmal subarachnoid hemorrhage in Glasgow coma score 3–5. Acta Neurochir (Wien). 2020;162(3):533–44. doi:10.1007/s00701-019-04190-y PubMed PMID: 31980948.

33. Brawanski N, Dubinski D, Bruder M, Berkefeld J, Hattingen E, Senft C, et al. Poor grade subarachnoid hemorrhage: Treatment decisions and timing influence outcome. Should we, and when should we treat these patients? Brain Hemorrhages. 2021 Mar;2(1):29–33. doi:10.1016/j.hest.2020.09.003

34. Autio AH, Paavola J, Tervonen J, Lång M, Huuskonen TJ, Huttunen J, et al. Clinical condition of 120 patients alive at 3 years after poor-grade aneurysmal subarachnoid hemorrhage. Acta Neurochir (Wien). 2021 Apr;163(4):1153–66. doi:10.1007/s00701-021-04725-2

35. Henry J, Dablouk MO, Kapoor D, Koustais S, Corr P, Nolan D, et al. Outcomes following poor-grade aneurysmal subarachnoid haemorrhage: a prospective observational study. Acta Neurochir (Wien). 2023;165(12):3651–64. doi:10.1007/s00701-023-05884-0 PubMed PMID: 37968366.

36. Ohbuchi H, Kasuya H, Hagiwara S, Kanazawa R, Yokosako S, Arai N, et al. Appropriate treatment within 13 hours after onset may improve outcome in patients with high-grade aneurysmal subarachnoid hemorrhage. Clin Neurol Neurosurg. 2023 Jul;230:107776. doi:10.1016/j.clineuro.2023.107776

37. Dablouk L, Dablouk MO, O’Sullivan MGJ. Outcomes and costs in the management of poor grade subarachnoid haemorrhage. Neurosurg Rev. 2025 Sep 24;48(1):663. doi:10.1007/s10143-025-03838-x

38. Fluss R, Fortunel A, Khatri D. Clinical outcome of high-grade aneurysmal acute subarachnoid hemorrhage: Hunt and Hess grade 4 survivors may have a good long-term functional outcome. Interdiscip Neurosurg. 2025;41:102095. doi:10.1016/j.inat.2025.102095

39. Alexandre AM, Caricato A, Pedicelli A, Marchese E, Scarcia L, Feletti A, et al. Delayed cerebral infarction in poor grade subarachnoid hemorrhage. Features, predictors, and clinical impact. Neurosurg Rev. 2025 Aug 26;48(1):620. doi:10.1007/s10143-025-03762-0

40. Goldberg J, Schoeni D, Mordasini P, Z’Graggen W, Gralla J, Raabe A, et al. Survival and Outcome After Poor-Grade Aneurysmal Subarachnoid Hemorrhage in Elderly Patients. Stroke. 2018 Dec;49(12):2883–9. doi:10.1161/STROKEAHA.118.022869

41. Hijdra A, Braakman R, Gijn J van, Vermeulen M, Crevel H van. Aneurysmal subarachnoid hemorrhage. Complications and outcome in a hospital population. Stroke. 1987 Nov;18(6):1061–7. doi:10.1161/01.str.18.6.1061

42. Ross N, Hutchinson PJ, Seeley H, Kirkpatrick PJ. Timing of surgery for supratentorial aneurysmal subarachnoid haemorrhage: report of a prospective study. J Neurol Neurosurg Psychiatry. 2002 Apr;72(4):480–4. PubMed PMID: 11909907; PubMed Central PMCID: PMC1737846.

43. Hutchinson PJ, Power DM, Tripathi P, Kirkpatrick PJ. Outcome from poor grade aneurysmal subarachnoid haemorrhage–which poor grade subarachnoid haemorrhage patients benefit from aneurysm clipping? Br J Neurosurg. 2000 Apr;14(2):105–9. doi:10.1080/02688690050004516 PubMed PMID: 10889881.

44. Wilby MJ, Sharp M, Whitfield PC, Hutchinson PJ, Menon DK, Kirkpatrick PJ. Cost-Effective Outcome for Treating Poor-Grade Subarachnoid Hemorrhage. Stroke. 2003 Oct;34(10):2508– 11. doi:10.1161/01.str.0000089922.94684.13 PubMed PMID: 12958321.

45. Laidlaw JD, Siu KH. Poor-grade Aneurysmal Subarachnoid Hemorrhage: Outcome after Treatment with Urgent Surgery. Neurosurgery. 2003 Dec;53(6):1275–82. doi:10.1227/01.neu.0000093199.74960.ff PubMed PMID: 14633294.

46. Mocco J, Ransom ER, Komotar RJ, Schmidt JM, Sciacca RR, Mayer SA, et al. Preoperative Prediction of Long-term Outcome in Poor-grade Aneurysmal Subarachnoid Hemorrhage. Neurosurgery. 2006 Sep;59(3):529–38. doi:10.1227/01.neu.0000228680.22550.a2 PubMed PMID: 17029345.

47. Kranthi S, Sahu B, Aniruddh P. Factors affecting outcome in poor grade subarachnoid haemorrhage: An institutional study. Asian J Neurosurg. 2016;11(04):365–71. doi:10.4103/1793-5482.149991 PubMed PMID: 27695539; PubMed Central PMCID: PMC4974960.

48. Zheng K, Zhao B, Tan XX, Li ZQ, Xiong Y, Zhong M, et al. Comparison of Aggressive Surgical Treatment and Palliative Treatment in Elderly Patients with Poor-Grade Intracranial Aneurysmal Subarachnoid Hemorrhage. BioMed Res Int. 2018;2018(1):5818937. doi:10.1155/2018/5818937 PubMed PMID: 29998135; PubMed Central PMCID: PMC5994574.

49. Panni P, Colombo E, Donofrio CA, Barzaghi LR, Albano L, Righi C, et al. Hemorrhagic burden in poor-grade aneurysmal subarachnoid hemorrhage: a volumetric analysis of different bleeding distributions. Acta Neurochir (Wien). 2019;161(4):791–7. doi:10.1007/s00701-019-03846-z PubMed PMID: 30790092.

50. Wang B, Li T, Zhao Y, Zhou T, Wang R, Li Y, et al. Long-term outcomes of poor-grade aneurysmal subarachnoid hemorrhage: a multicenter observational cohort study. J Neurosurg. 2026 Feb 1;1–10. doi:10.3171/2025.10.JNS25978

51. Petruk KC, West M, Mohr G, Weir BK, Benoit BG, Gentili F, et al. Nimodipine treatment in poor-grade aneurysm patients. Results of a multicenter double-blind placebo-controlled trial. J Neurosurg. 1988 Apr;68(4):505–17. doi:10.3171/jns.1988.68.4.0505 PubMed PMID: 3280746.

52. De Oliveira Manoel AL, Mansur A, Murphy A, Turkel-Parrella D, Macdonald M, Macdonald RL, et al. Aneurysmal subarachnoid haemorrhage from a neuroimaging perspective. Crit Care. 2014 Nov 13;18(6):557. doi:10.1186/s13054-014-0557-2

53. McNeill L, English SW, Borg N, Matta BF, Menon DK. Effects of Institutional Caseload of Subarachnoid Hemorrhage on Mortality: A Secondary Analysis of Administrative Data. Stroke. 2013 Mar;44(3):647–52. doi:10.1161/STROKEAHA.112.681254

54. Bardach NS, Olson SJ, Elkins JS, Smith WS, Lawton MT, Johnston SC. Regionalization of Treatment for Subarachnoid Hemorrhage: A Cost-Utility Analysis. Circulation. 2004 May 11;109(18):2207–12. doi:10.1161/01.CIR.0000126433.12527.E6

55. Yang Y, Wang B, Ge X, Lu W, Peng C, Zhao Y, et al. Natural Course of Ruptured but Untreated Intracranial Aneurysms: A Multicenter 2-Year Follow-Up Study. Stroke. 2023 Aug;54(8):2087–95. doi:10.1161/STROKEAHA.123.042530

56. the Unruptured Intracranial Aneurysms and SAH CDE Project Investigators, Suarez JI, Sheikh MK, Macdonald RL, Amin-Hanjani S, Brown RD, et al. Common Data Elements for Unruptured Intracranial Aneurysms and Subarachnoid Hemorrhage Clinical Research: A National Institute for Neurological Disorders and Stroke and National Library of Medicine Project. Neurocrit Care. 2019 Jun;30(S1):4–19. doi:10.1007/s12028-019-00723-6

57. Al-Khindi T, Macdonald RL, Schweizer TA. Cognitive and Functional Outcome After Aneurysmal Subarachnoid Hemorrhage. Stroke. 2010 Aug;41(8). doi:10.1161/STROKEAHA.110.581975

